# Effect of low dose naltrexone for long covid: a systematic review

**DOI:** 10.1101/2025.09.09.25335451

**Authors:** Oyungerel Byambasuren, Tiffany Atkins, Shaira Baptista, Paul Glasziou, Samantha Chakraborty

## Abstract

**Background:** Long covid is a debilitating chronic condition, and the effect of low dose naltrexone (LDN) on its symptoms is unclear. We aimed to determine the effectiveness of LDN on symptoms of long covid.

**Methods:** We searched PubMed, Embase, and Cochrane Library for published studies; ClinicalTrials.gov and World Health Organization International Clinical Trials Registry Platform for registered ongoing studies from inception to 1 May 2025. Eligible studies were randomised controlled trials or pre-post studies in patients with long covid reporting on fatigue, quality of life, cognition, or other symptoms. Risk of bias was assessed by Newcastle-Ottawa scale.

**Results:** Of 226 titles and abstracts screened, no randomised controlled trials were identified. Four observational pre-post studies from USA and Ireland (n=155) met inclusion criteria. LDN doses varied from 1mg/d to 6 mg/d. Pooled analyses showed moderate effects for reducing fatigue (Hedges’ g= -0.74; 95% CI [-1.11, -0.37]; p<0.001), brain fog (Hedges’ g= - 0.53; 95%CI [-1.01, -0.05]; p=0.03), and improving sleep quality (Hedges’ g= -0.60; 95%CI [-0.91, -0.30]; p=0.0001), and large effects for pain (Hedges’ g= -0.93; 95%CI [-1.29, -0.57]; p<0.001) and daily functioning (Hedges’ g= -0.93; 95%CI [-1.29, -0.57]; p<0.0001) in favour of LDN. Heterogeneity ranged from 0% to 62%. Risk of bias was assessed as low in all four studies. No serious adverse events were reported in the two studies that assessed safety.

**Conclusion:** Limited evidence from small pre-post studies suggests LDN may improve fatigue, cognition, sleep, pain, and functioning in long covid. However, certainty of evidence is low. Well-powered trials are urgently needed to confirm efficacy, determine dosing and duration, and identify subgroups most likely to benefit.

**Protocol registration:** Open Science Framework https://doi.org/10.17605/OSF.IO/C2VKX

## Introduction

The COVID-19 pandemic has given rise to a novel chronic condition known as Post-Acute Sequelae of COVID-19 or long covid. This condition is characterised by persistent symptoms such as fatigue, respiratory issues, and cognitive impairments, which can cause prolonged illness and disability (1, 2). Conservative estimates suggest that around 10% of people infected with COVID-19 develop long covid, translating to millions of cases worldwide (3).

Among the plethora of long covid symptoms, the most debilitating are fatigue, post-exertional malaise, and cognitive dysfunction (4). These symptoms have profound personal, social, psychological, and financial consequences (5). There is an urgent need for effective, evidence-based treatment options for long covid.

There is a substantial symptom overlap between Myalgic Encephalomyelitis/Chronic Fatigue Syndrome (ME/CFS) and long covid. Both conditions are also associated with impairment of the TRPM3 ion channel, pointing to a potential common underlying biological mechanism and highlighting TRPM3 as a therapeutic target (6, 7).

One proposed off-label treatment for this impairment is low dose naltrexone (LDN). Naltrexone in standard doses (50-100mg/day) is approved as an opioid antagonist for treating alcohol use disorder and opioid dependence (6, 7). In contrast, much lower doses (∼4.5mg/day) have been used to manage chronic pain, ME/CFS, and other chronic neuroimmune disorders such as Crohn’s disease and fibromyalgia, due to its biological plausibility and favourable safety profile (7-10).

Although clinical effectiveness studies are scarce, off-label use of LDN is increasing (11, 12). This review therefore aims to systematically summarise all available evidence on the effectiveness of LDN for alleviating symptoms of long covid.

## Methods

We conducted a systematic review using enhanced processes and automation tools and reported it according to the Preferred Reporting Items for Systematic Reviews and Meta-Analyses (PRISMA) statement (13). Our protocol was pre-registered on the Open Science Framework (OSF) (https://doi.org/10.17605/OSF.IO/C2VKX). Ethics approval was not required.

We searched PROSPERO and OSF databases to confirm no similar reviews were underway. We then searched PubMed, Embase (Elsevier) and the Cochrane Library for published studies, ClinicalTrials.gov and World Health Organization (WHO) - International Clinical Trials Registry Platform (ICTRP) for registered ongoing studies from inception to 1 May 2025.

Search strings combined MeSH or other subject terms, synonyms and search filters, and were designed with the Systematic Review Accelerator suite of tools (14). An information specialist peer-reviewed the strategy using PRESS guidelines (see Supplement 1). No restrictions were applied for language or publication type. We performed backward and forward citation searching using SpiderCite (14).

### Eligibility Criteria

We included randomised controlled trials and pre-post cohort studies enrolling patients with long COVID, defined according to the WHO clinical case definition, where the majority of participants were at least 12 weeks post-infection (15). Case studies and case series were excluded. Studies conducted in either primary or secondary care settings were eligible.

□ Participants: Adults with long covid (with the majority at least 12 weeks post-covid or with a reported subset) were eligible for this review. Studies focusing on acute and post-acute covid (within 12 weeks of the onset of COVID-19 symptoms) were excluded.
□ Interventions: low dose naltrexone (1-10mg) as sole or add-on intervention.
□ Comparator: usual care or placebo.
□ Outcomes: Primary outcomes were fatigue and/or quality of life. Secondary outcomes included cognitive symptoms or function, other long covid symptoms, and adverse events (e.g., gastrointestinal symptoms). We also noted any additional outcomes reported.

### Study selection and screening

Two authors (OB, SC) independently screened titles and abstracts. Full texts of potentially eligible articles were retrieved and assessed by OB and SC. Citation and trial registry searches were screened by OB and TA. Disagreements were resolved by discussion or consultation with all authors.

### Data extraction

We piloted a standardised form on three studies and then extracted data in duplicate (TA, SB). Extracted items included:

□ Study details: study authors, year, country, study design, duration of follow-up, setting
□ Participant characteristics: sample size, age, gender, time since COVID-19 infection
□ Intervention details: dose, frequency and comparator/s
□ Reported outcomes: fatigue, quality of life, cognitive symptoms or function, other covid symptoms, adverse events

### Risk of bias assessment

We planned to use the Cochrane Risk of Bias 2.0 tool for RCTs and the Newcastle–Ottawa Scale (NOS) for observational studies (16). Two authors (TA, SB) independently assessed risk of bias. For pre–post designs, we adapted the NOS by removing non-applicable subsections and adjusting the scoring accordingly.

### Data analysis

We calculated Hedges’ g effect sizes for pre–post differences. Interpretation thresholds were: >−0.20 = negligible effect; −0.20 to −0.49 = small; −0.50 to −0.79 = moderate; ≥−0.80 = large (17). Where multiple similar outcomes were reported (e.g., different fatigue measures), we selected the outcome most common across studies to allow pooling to ensure only one outcome per study contributed. Outcomes that could not be combined in a meta-analysis were described narratively.

We pooled effect sizes using a random-effects model (DerSimonian–Laird) and assessed heterogeneity with the I^2^ statistic. We searched trial registries for unpublished studies but did not assess publication bias because fewer than 10 studies were eligible. Planned subgroup analyses (baseline severity, LDN dose) were not feasible due to insufficient data.

All analyses were conducted using *Stata* version 16.1/MP (StataCorp, 2019).

## Results

Of 226 records screened, four full-text articles met the inclusion criteria (Figure 1). All included studies were pre-post cohort designs, conducted in the United States (n=3) and Ireland (n=1), and together included 155 patients (18-21) (Table 1). LDN was used as a standalone intervention in three studies; one study (Isman 2024(19)) combined it with nicotinamide adenine dinucleotide (NAD+). Two studies reported minor adverse events (19, 20). No RCTs were identified, but three registered trials were ongoing at the time of this review with no published results available.

**Table 1.** Characteristics of included studies.

| Study ID, location | Study type, timeframe | Population | Intervention | Outcomes | Adverse events |
| --- | --- | --- | --- | --- | --- |
| **Bonilla (18) 2023**<br>USA | Retrospective cohort, 18 May 2021 - 18 Mar 2023 | N=59, median age 45, 68% female, median duration of symptoms 361 days | Individualised LDN dose-titration ranging from 0.5mg/d to 6mg/d for median of 143 days | Total number of symptoms, severity in subset of symptoms (fatigue, PEM, unrefreshing sleep, and abnormal sleep pattern) and FSS | No information provided |
| **Isman (19) 2024**<br>USA | Prospective cohort, 31 Mar 2021 to 22 Dec 2022 | N=36, mean age 45 (28-69), 70% female, mean duration of symptoms 223 days | LDN titrated from 1mg/d for 4 days, 2mg/d for 4 days and 4.5 mg/d thereafter, 400 mg/mL NAD+ was applied via iontophoresis patches weekly for 12 weeks | Fatigue, pain, and quality of life, | Mild adverse events (skin irritation at location of NAD+ patch, nausea, fatigue, dizziness, and low mood) |
| **O'Kelly (20) 2022**<br>Ireland | Prospective cohort, Jun – Nov 2020 | N=36, median age 44 yrs, 77% female, median duration of symptoms 333 days. | LDN 1mg/d in first month, 2mg/d in second month, 3mg/d for third month. | Recovery from COVID-19, limitation in activities of daily living, energy levels, pain levels, levels of concentration and sleep disturbance | Two participants had same adverse events (diarrhoea and fatigue) |
| **Tamariz (21) 2024**<br>USA | Retrospective cohort, 2021 to Jan 2023 | N=24, mean age 54, 34% female, (duration of symptoms not reported) | LDN 1.5–4.5 mg/d, median duration of follow up 147 days | Improvement in fatigue, pain, brain fog, or dyspnoea at least 1 month after starting treatment | No information provided |

LDN - low dose naltrexone, PEM - post-exertional malaise, FSS - functional status scale, NAD - nicotinamide adenine dinucleotide

**Figure 1.**
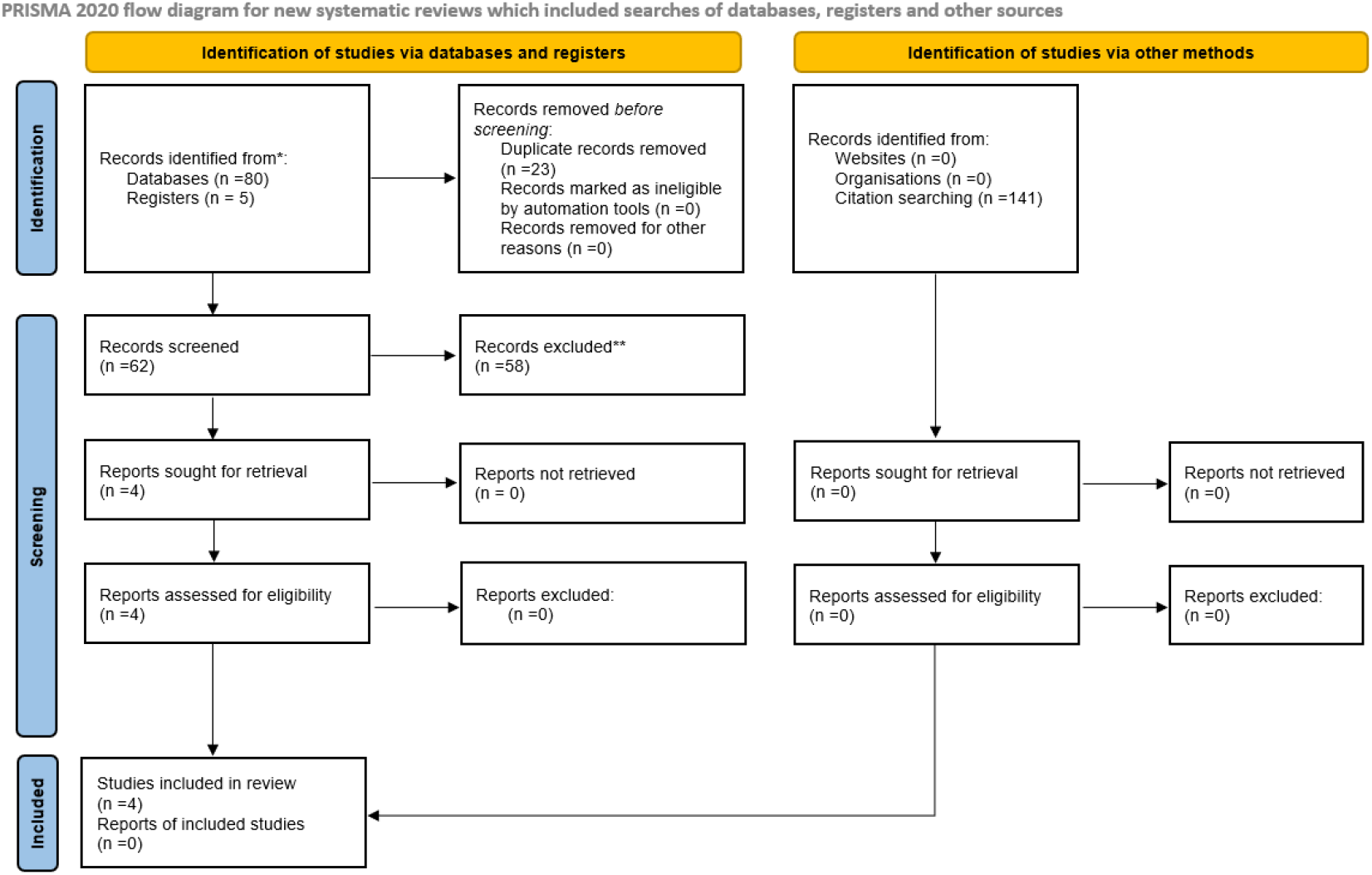
Screening and selection of studies.

Forest plots (Figures 2 to 6) summarise the effect sizes (Hedges’ g) from individual studies and the pooled effect sizes for each outcome. Scale directions were standardised so that negative values consistently favoured LDN. Figure 7 presents all effect sizes in a single combined plot. A detailed summary of outcomes is provided in Supplement 2.

**Figure 2.**
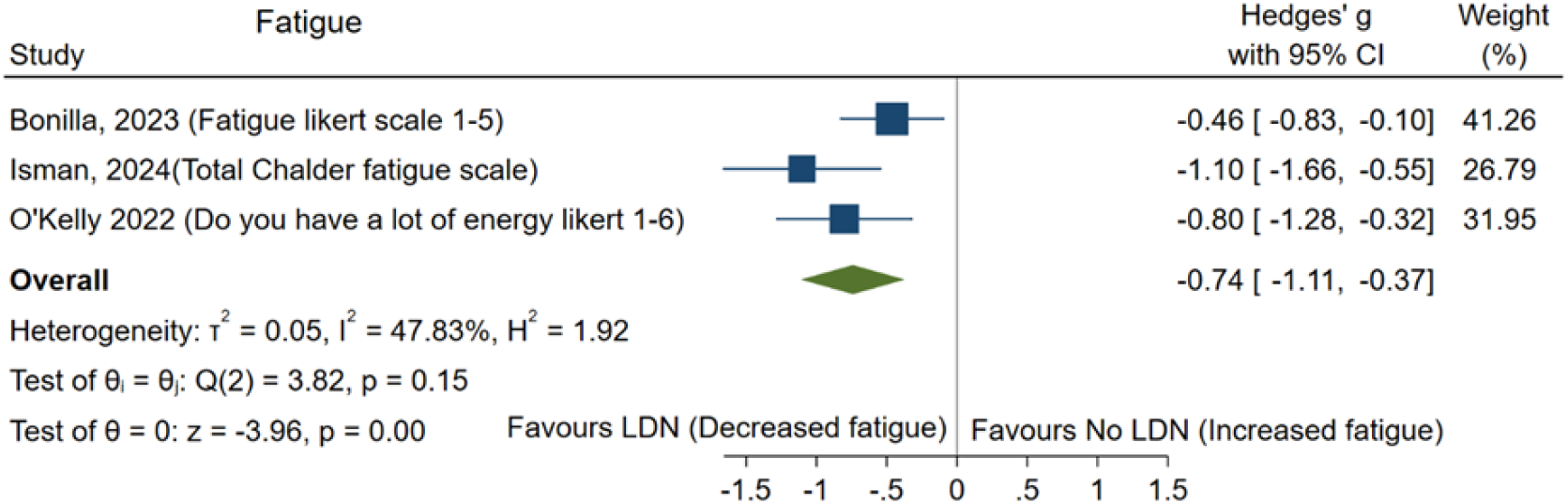
Forest plot of fatigue scores.

### Fatigue

Three studies reported fatigue outcomes, which were pooled (18-20). The meta-analysis showed a moderate effect size favouring LDN for reducing fatigue (Hedges’ g= -0.74; 95% CI [-1.11, -0.37]; p<0.001; I^2^=47.7%) (Figure 2). Two additional fatigue-related outcomes also favoured LDN: post exertional malaise from (Bonilla 2023(18); Hedges’ g=-0.48; 95% CI [-0.84 to -0.11]; p=0.010) and the SF 36 energy/fatigue subscale (Isman 2024(19); Hedges’ g=-1.10; 95%CI [-1.66 to -0.55]; p<0.0001) (Supplement 2). In Tamariz 2024, 13 of 24 patients (54.2%) reported improved fatigue after starting LDN (21).

### Pain

Two studies reported pain outcomes, which were pooled (19, 20). The meta-analysis showed a large effect size favouring LDN for reducing pain (Hedges’ g= -0.93; 95%CI [-1.29, -0.57]; p<0.001) with no heterogeneity (I^2^ = 0%) (Figure 3). In Tamariz 2024, 12 of 24 patients (50%) reported improvements in pain after starting LDN (21) (Supplement 2).

**Figure 3.**
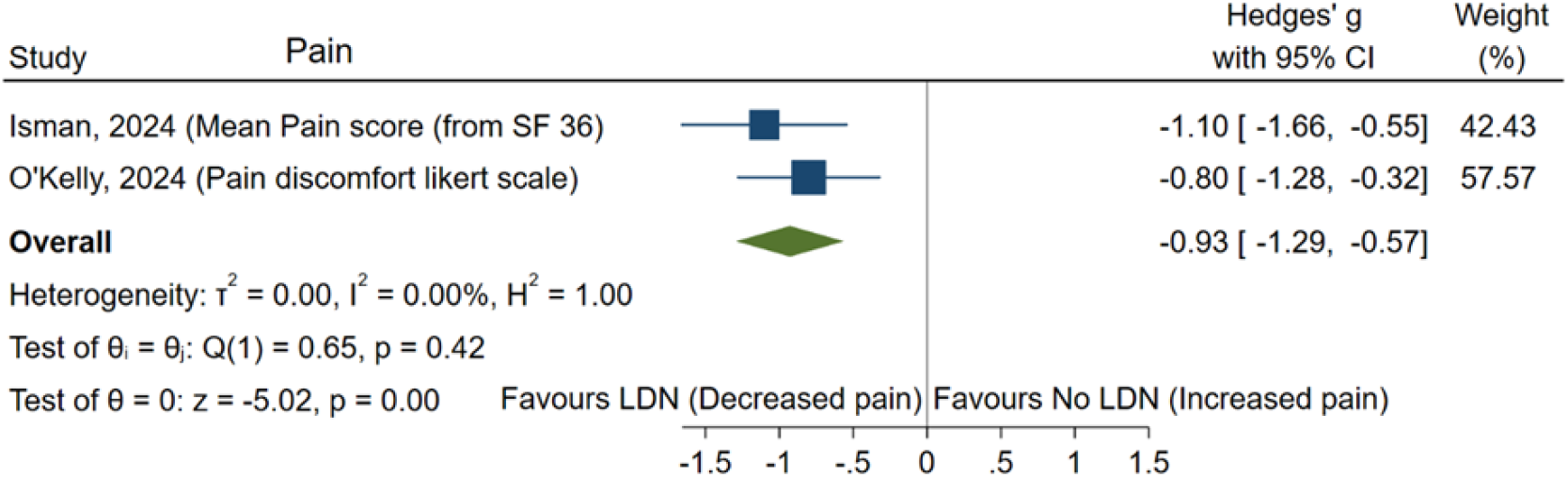
Forest plot of pain scores.

### Brain Fog

Two studies reported brain fog outcomes, which were pooled (18, 20). The meta-analysis showed a moderate effect size favouring LDN for reducing brain fog (Hedges’ g= -0.53; 95%CI [-1.01, -0.05]; p=0.03) (Figure 4) with moderate heterogeneity (I^2^ = 61.8%). In Tamariz 2024, 4 of 24 patients (16.7%) reported improvements in brain fog after starting LDN (21) (Supplement 2).

**Figure 4.**
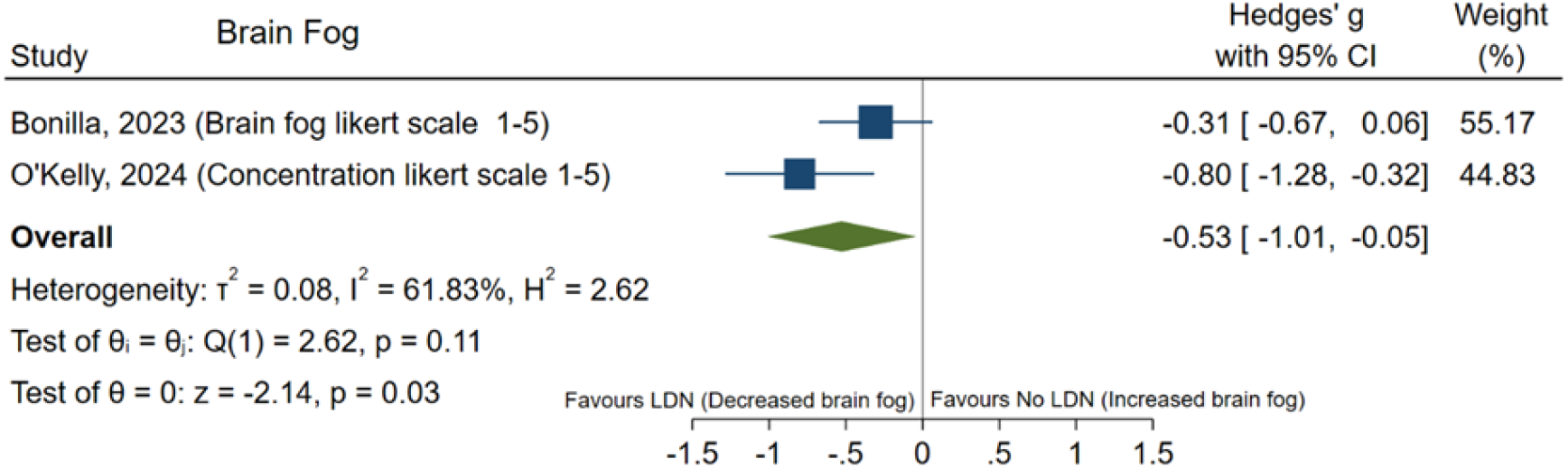
Forest plot of brain fog scores.

**Figure 5.**
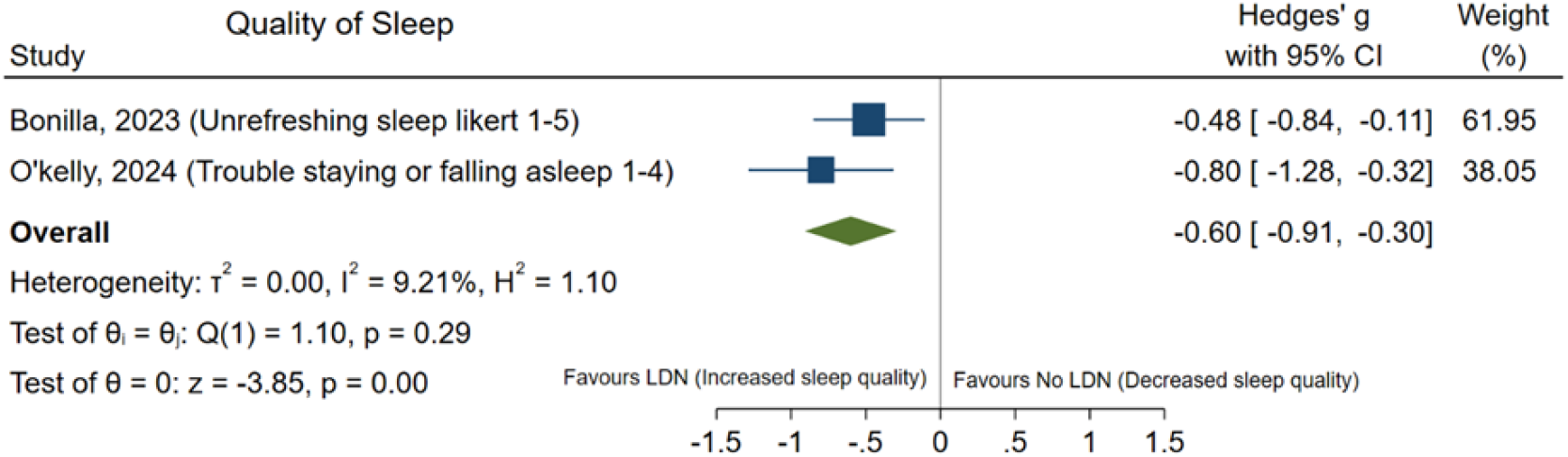
Forest plot of quality of sleep.

### Quality of Sleep

Two studies reported sleep quality outcomes, which were pooled (18, 20). The meta-analysis showed a moderate effect size favouring LDN for improving sleep quality (Hedges’ g= -0.60; 95%CI [-0.91, -0.30]; p=0.0001) with low heterogeneity (I^2^ = 9.2%). Additionally, Bonilla 2023 reported a small effect on reducing an abnormal sleep pattern favouring LDN (Hedges’ g= -0.45; 95% CI [-0.81 to -0.08]; p=0.016) (18) (Supplement 2).

### Daily Functioning

Two studies reported daily functioning outcomes, which were pooled (19, 20). The meta-analysis showed a large effect favouring LDN for improving daily functioning (Hedges’ g= - 0.93; 95%CI [-1.29, -0.57]; p<0.0001) with no heterogeneity (I^2^ = 0.0 %) (Figure 6).

**Figure 6.**
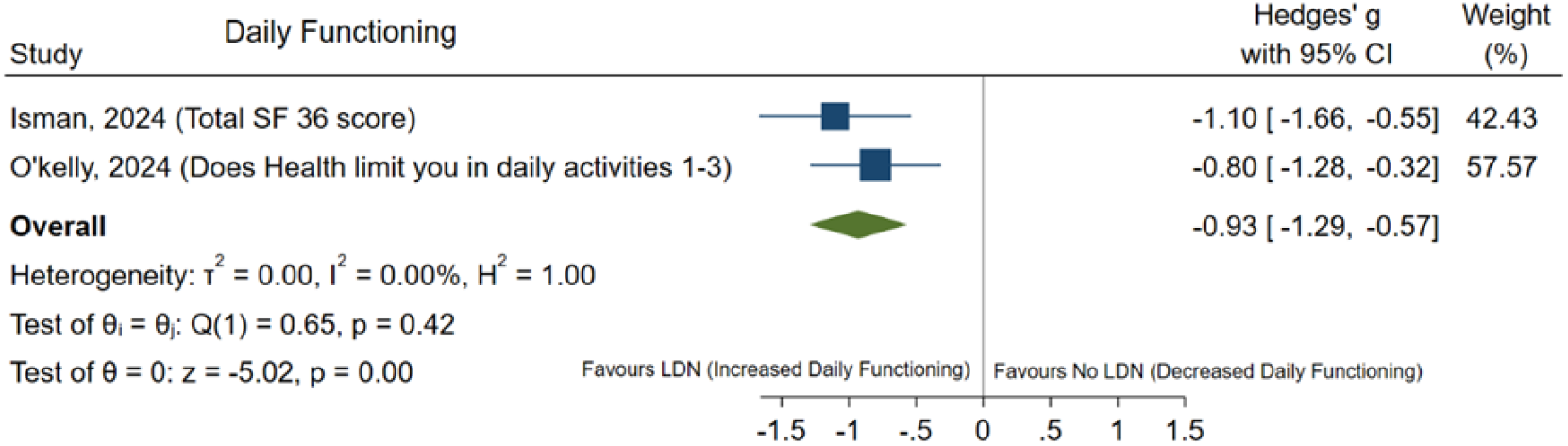
Forest plot of daily functioning scores.

**Figure 7.**
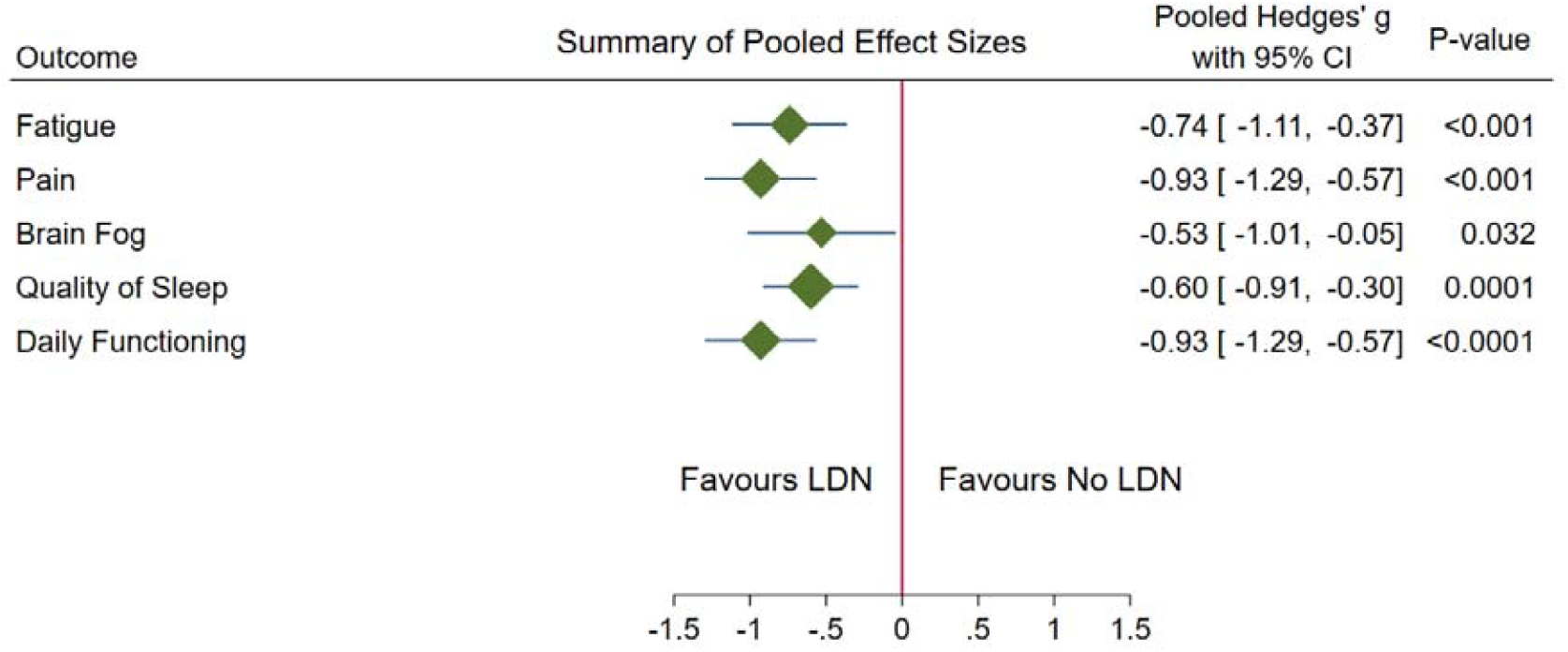
Summary of pooled effect sizes for all outcomes.

### Summary of pooled effects

Figure 7 presents the pooled effect sizes for all five outcomes. LDN demonstrated moderate to large effects across fatigue, pain, brain fog, sleep quality, and daily functioning.

## Risk of bias

Table 2 summarises the risk of bias for the included studies using the Newcastle-Ottawa scale (16). The maximum points for the Selection domain were reduced by two to account for the not applicability of “selection of the non-exposed cohort” and the “outcome of interest was not present at the start of the study”. The Comparability domain was reduced by one point due to the inapplicability of “the study controls for the most important factor”. Consequently, the maximum overall quality score was 6 points. All four studies scored either four or five points, indicating a low risk of bias.

**Table 2.** Risk of bias of included studies by Newcastle-Ottawa Scale.

| Assessment domains | Selection (Max 2 points) |  |  |  | Comparability (Max 1 points) | Outcome (Max 3 points) |  |  | Overall Quality Scores (Max 6 points) | Risk of bias level |
| --- | --- | --- | --- | --- | --- | --- | --- | --- | --- | --- |
|  | Representativeness of the Sample | Selection of Nonexposed Cohort | Ascertainment of Exposure | Outcome of Interest Was Not Present at Start of Study | Comparability Based on Design and Analysis | Assessment of the Outcome | Outcome Follow-up | Adequacy of Follow-up |  |  |
| **Bonilla 2023** | 1 | N/A | 1 | N/A | 1 | 0 | 1 | 1 | 5 | Low |
| **Isman 2024** | 1 | N/A | 1 | N/A | 1 | 0 | 1 | 0 | 4 | Low |
| **O’Kelly 2022** | 1 | N/A | 1 | N/A | 0 | 0 | 1 | 1 | 4 | Low |
| **Tamariz 2024** | 1 | N/A | 1 | N/A | 1 | 0 | 1 | 1 | 5 | Low |

### Registered trials

We identified three ongoing registered trials investigating LDN for long covid (Table 3). The first is a phase 2 double-blind placebo-controlled RCT in British Columbia, Canada, evaluating titrated doses of LDN for post-COVID-19 fatigue syndrome over 16 weeks, with a target enrolment of 160 participants and an expected completion date of Dec 2024 (22, 23). The second is a phase-1 trial in Australia testing titrated dose of LDN for post-COVID-19 condition over 12 weeks, with a planned recruitment of 56 participants (24). The third, also led by the same Australian team, is assessing LDN for both ME/CFS and long covid (25).

**Table 3.** Summary of registered trials (n=3)

| Registration ID | Status | Intervention & dose | Sample size | Treatment Duration | Primary Outcome |
| --- | --- | --- | --- | --- | --- |
| NCT05430152<br>(22, 23) | Recruiting | LDN as a compounded capsule starting at 1 mg/day and increasing up to 4.5 mg/day (by week 4) | **N=160** | 16 weeks | Change in fatigue intensity by 4.7 points over using the Fatigue Severity Scale (FSS) |
| ACTRN<br>12623001042639<br>(24)<br>NALCOVID study | Recruiting | LDN 3-6mg/day | **N=56** | 12 weeks | Detectable change in symptom presentation and severity measured by DSQ Symptom Inventory Questionnaire |
| ACTRN<br>12624001162505<br>(25)<br>TreatMELC study | Not yet recruiting | LDN start at 1.5mg/day and increase by 1.5mg/day weekly until their maximum dose is reached (target 4-6mg/day) | **N=56** | 12 weeks | Change in the Transient receptor potential cation channel subfamily M member 3 (TRPM3) function |

## Discussion

### Principal findings

We found no randomised controlled trials, but four observational studies reported outcomes of LDN for long covid. Collectively, these studies demonstrated moderate to large improvements from pre-to post-treatment measurements in fatigue, brain fog, pain, sleep quality, and daily functioning, with standardised mean difference ranging from 0.53 for brain fog to 0.93 daily functioning. No serious adverse events were reported. All four studies were non-randomised, and most were assessed as having low risk of bias, but overall certainty of evidence remains low.

### Strengths and limitations

The strengths of our review include a comprehensive search of multiple databases for both published and unpublished studies and a critical risk of bias assessment to identify potential sources of bias. However, the uncontrolled design of included studies precludes causal inference. These results have several limitations in particular the observed improvements could reflect the natural history of the illness, regression to the mean, or placebo effects, as all outcomes were self-reported. Publication bias is also a concern, since negative studies are less likely to be published and are not required to be registered, hence we cannot use registries to check for non-publication of known studies. Another limitation was the heterogeneity of outcome measures used across studies, which complicate pooling and interpretation of results.

### Interpretation and comparison with other evidence

The effect sizes observed in this review are comparable to or greater than those reported for interventions trialled in ME/CFS and fibromyalgia, conditions that share symptomatic and mechanistic overlap with long covid (9, 10). For example, pharmacological agents used for chronic fatigue and chronic pain often achieve only small improvements in symptom scores. The magnitude of change associated with LDN, if replicated in controlled trials, could therefore represent a clinically meaningful advance. Mechanistically, LDN may reduce neuroinflammation through modulation of microglial activation and antagonism of Toll-like receptor 4, and may restore TRPM3 ion channel function, which is impaired in both long covid and ME/CFS. These plausible biological pathways lend further weight to the observed clinical improvements.

### Implications for current practice and future research

LDN has been proposed for ME/CFS and other chronic inflammatory conditions, with case reports and series suggesting possible benefit, but controlled trials remain lacking for long covid. Despite this limited evidence, two recent clinical guides on infection-associated chronic illness management that included long covid recommended LDN as a potential treatment for fatigue, pain and post-exertional malaise (26, 27). Its low cost, oral administration, and established safety profile make it an attractive candidate for repurposing.

However, accessibility is currently limited because LDN must be compounded into low dose preparations and many clinicians are reluctant to prescribe it off-label. If robust evidence of effectiveness emerges, these barriers could be addressed, making LDN a low-cost treatment option that reduces financial burden for patients and expands access to care. In that scenario, LDN could provide a more feasible and equitable option than higher-cost alternatives such as antivirals or biologics, particularly in resource-limited settings where treatment choices for long COVID are scarce.

We identified three ongoing trials testing LDN in long covid, two of which are randomised and controlled. These studies will be crucial in confirming efficacy, clarifying optimal dosing and titration schedules, and establishing treatment duration. Future trials should incorporate validated patient-reported outcomes as well as objective measures of fatigue, cognitive function, and physical activity. Stratification by symptom phenotype and comorbidities will also be important to identify subgroups most likely to benefit.

Together, these considerations underscore the urgent need for well-designed, high-quality trials to establish whether LDN can be safely and effectively integrated into long COVID management.

## Conclusion

Preliminary observational evidence suggests LDN may improve fatigue, brain fog, pain, sleep quality, and daily functioning in patients with long COVID. However, the evidence summarised here is of low certainty. A well-powered randomised controlled trial is urgently needed to establish efficacy, determine optimal use, and confirm safety in this population. Given the global burden of long COVID and the accessibility of LDN, high-quality research is warranted to clarify its role in clinical practice.

## Supporting information

Supplement 1. Full search strategy

Supplement 2. Outcomes of individual studies

## Data Availability

All data produced in the present work are contained in the manuscript and the supplementary material.

## Author contributions

PG, OB, and SC conceived the study and co-designed the study with TA and SB. TA led the literature searches. OB and TA conducted parallel title, abstract, and full text screening. OB and TA were responsible for data extraction and formal analysis. TA and SB did risk of bias assessments of included studies. All authors contributed to resolving disagreements throughout the study and to writing of the manuscript.

## Ethics approval

Not applicable.

