## Supplement 1. Full search strategy for "Effect of low dose naltrexone for long covid: a systematic review"

Supplement 1 - Search Strategies

**PubMed - 01-05-2025 returned n=16 results**

("Post-Acute COVID-19 Syndrome"[Mesh] OR "COVID-19/complications"[Majr] OR "post covid"[tiab] OR "post-covid"[tiab] OR postcovid[tiab] OR "post coronavirus"[tiab] OR postcoronavirus[tiab] OR "long covid"[tiab]  OR "Long-covid"[tiab] OR “COVID-19 survivors”[tiab] OR “COVID-19 related”[tiab] OR “history of COVID-19”[tiab] OR “history of COVID”[tiab] OR “Subacute COVID-19”[tiab] OR “after COVID-19”[tiab] OR “subacute COVID-19”[tiab] OR “after covid”[tiab] OR "After SARS"[tiab] OR "Post-Acute Coronavirus"[tiab] OR PASC[tiab] OR ((Post-discharge[tiab]) AND ("Covid-19"[ti])) OR (History[tiab] AND "COVID-19 infection"[tiab]))

AND

(“Naltrexone”[Mesh] OR “Naltrexone”[tiab] OR “low-dose Naltrexone” [tiab])

NOT

(Animals[Mesh] NOT (Animals[Mesh] AND Humans[Mesh]))

**Cochrane - 01-05-2025 returned n=8**

([mh "Post Acute COVID 19 Syndrome"] OR [mh "COVID 19"/CO] OR "post covid":ti,ab OR post-covid:ti,ab OR postcovid:ti,ab OR "post coronavirus":ti,ab OR postcoronavirus:ti,ab OR "long covid":ti,ab OR Long-covid:ti,ab OR "COVID-19 survivors":ti,ab OR "COVID-19 related":ti,ab OR "history of COVID-19":ti,ab OR "history of COVID":ti,ab OR "Subacute COVID-19":ti,ab OR "after COVID-19":ti,ab OR "subacute COVID-19":ti,ab OR "after covid":ti,ab OR "After SARS":ti,ab OR "Post-Acute Coronavirus":ti,ab OR PASC:ti,ab OR ((Post-discharge:ti,ab) AND (Covid-19:ti)) OR (History:ti,ab AND "COVID-19 infection":ti,ab))

AND
([mh Naltrexone] OR Naltrexone:ti,ab OR "low-dose Naltrexone":ti,ab)

**Embase - 01-05-2025 returned n=56**

('long COVID'/exp/mj OR 'post covid':ti,ab OR post-covid:ti,ab OR postcovid:ti,ab OR 'post coronavirus':ti,ab OR postcoronavirus:ti,ab OR 'long covid':ti,ab OR Long-covid:ti,ab OR 'COVID-19 survivors':ti,ab OR 'COVID-19 related':ti,ab OR 'history of COVID-19':ti,ab OR 'history of COVID':ti,ab OR 'Subacute COVID-19':ti,ab OR 'after COVID-19':ti,ab OR 'subacute COVID-19':ti,ab OR 'after covid':ti,ab OR 'After SARS':ti,ab OR 'Post-Acute Coronavirus':ti,ab OR PASC:ti,ab OR ((Post-discharge:ti,ab) AND (Covid-19:ti)) OR (History:ti,ab AND 'COVID-19 infection':ti,ab))

AND
(Naltrexone/exp OR Naltrexone:ti,ab OR 'low-dose Naltrexone':ti,ab)

NOT ('animal'/exp NOT ('animal'/exp AND 'human'/exp))

**ClinicalTrials.gov**

condition; Post COVID condition; other terms; Long covid, Intervention/treatment as Low-dose Naltrexone

**World Health Organization – International Clinical Trials Registry Platform (ICTRP**)

advanced search; in Title; Long covid OR post covid condition OR naltrexone, in the condition; Long covid OR Post covid condition; in the intervention, AND Low-dose Naltrexone
