## Supplement 2. Outcomes of individual studies for "Effect of low dose naltrexone for long covid: a systematic review"

**Supplement 2. Summary of outcomes of individual studies**

|  |  | **Before** | | **After** | |  |  |
| --- | --- | --- | --- | --- | --- | --- | --- |
| **Study ID** | **Measurement tool** | **n** | **Mean (SD)** | **n** | **Mean (SD)** | **P value** | **Effect size**  **Hedges’ g (95%CI)** |
| FATIGUE |  |  |  |  |  |  |  |
| Bonilla, 2023 | *Fatigue likert scale (1-5) | 59 | 3.97 (1.08) | 59 | 3.41 (1.59) | 0.013 | -0.46  (-0.83 to -0.10) |
| Bonilla, 2023 | *Post exertional malaise  (1-5) | 59 | 3.76 (1.37) | 59 | 3.14 (1.67) | 0.010 | -0.48  (-0.84 to -0.11) |
| Isman, 2024 | #Mean energy/fatigue (from SF 36) | 36 | 14.7(11.3) | 23 | 35.9(26.3) | <0.0001 | -1.10  (-1.66 to -0.55) |
| Isman, 2024 | *Total sum of chalder fatigue scale | 36 | 25.9(4.6) | 23 | 17.4(9.7) | <0.0001 | -1.10  (-1.66 to -0.55) |
| O’Kelly, 2022 | #Do you have a lot of energy likert (1-6) | 36 | Median (IQR) 3(2-3) | 36 | Median (IQR)  3(3-4) | 0.001 | -0.80  (-1.28 to -0.32) |
| Tamariz, 2024 | Reported improvements in fatigue n (%) | 24 |  | 24 | n=13(54.2%) |  |  |
| PAIN |  |  |  |  |  |  |  |
| Isman, 2024 | #Mean Pain score (from SF 36) | 36 | 47.5 (25.2) | 23 | 63.8 (29.9) | <0.0001 | -1.10  (-1.66 to -0.55) |
| O’Kelly, 2022 | #Pain discomfort likert scale (1-5) | 36 | Median (IQR) 2(2-3) | 36 | Median (IQR)  4(3-4) | <0.001 | -0.80 (-1.28 to -0.32) |
| Tamariz, 2024 | Reported improvements in Pain, n (%) | 24 |  | 24 | n=12 (50.0%) |  |  |
| BRAIN FOG | | | | | | | |
| Bonilla, 2023 | *Brain fog likert scale (1-5) | 59 | 3.14 (1.59) | 59 | 2.78 (1.88) | 0.097 | -0.31  (-0.67 to 0.06) |
| O’Kelly, 2022 | #Concentration likert scale (1-5) | 36 | Median (IQR)  2(1-2) | 36 | Median (IQR)  2(2-3) | 0.001 | -0.80 (-1.28 to -0.32) |
| Tamariz, 2024 | Reported improvements in Brain Fog n (%) | 24 |  | 24 | n=4 (16.7%) |  |  |
| QUALITY OF SLEEP | | | | | | | |
| Bonilla, 2023 | #Unrefreshing sleep likert (1-5) | 59 | 3.48 (1.54) | 59 | 2.85 (1.86) | 0.010 | -0.48  (-0.84 to -0.11) |
| Bonilla, 2023 | #Abnormal sleep pattern likert  (1-5) | 59 | 2.76 (2.00) | 59 | 2.05 (1.88) | 0.016 | -0.45  (-0.81 to -0.08) |
| O’Kelly, 2022 | *Trouble staying or falling asleep (1-4) | 36 | Median (IQR)  2 (1-3) | 36 | Median (IQR)  3 (1-3) | <0.001 | -0.80  (-1.28 to -0.32) |
| DAILY FUNCTIONING | | | | | | | |
| O’Kelly, 2022 | *Does Health limit you in daily activities (1-3) | 36 | Median (IQR)  1 (1-2) | 36 | Median (IQR)  2 (1-2) | 0.001 | -.80  (-1.28 to -0.32) |
| Isman, 2024 | *Total SF 36 score | 36 | 36.5(15.6) | 23 | 52.1(24.8) | <0.0001 | -1.10  (-1.66 to -0.55) |

*> score > fatigue/pain/brain fog/quality of sleep/daily functioning

\# > score < fatigue/pain/brain fog/quality of sleep/daily functioning

^H^ When calculating the Hedges’ g, different scale directions were converted to the same direction for consistency. Hedges’ g: - values favour LDN and +values do not favour LDN.
